# NMF Deconvolution of a High-ROS Transcriptional Program Uncovers mTOR-Dependent Therapeutic Sensitivity in Stomach Adenocarcinoma

**DOI:** 10.64898/2026.04.12.26350699

**Authors:** Rishav Roy, Jayasree Patnaik, Averi Chakraborty, Srinivas Patnaik, Tithi Parija

## Abstract

**Background:** Stomach adenocarcinoma is driven by heterogeneity, limiting therapeutic success. Although ROS acts as a continuous “redox rheostat” for tumor evolution, it is categorized based on binary models that are masked by tumor-microenvironment (TME) confounders. Here, we have defined a continuous, TME-independent ROS axis to help identify intrinsic vulnerabilities and improve patient stratification.

**Methods:** Non-negative matrix factorization (NMF) defined a ROS-Axis in TCGA-STAD which was validated in ACRG Cohort. Multivariate regression model isolated intrinsic signatures via “residual” ROS scores by adjusting for TME confounders. Survival was assessed using Cox hazard models. Drug sensitivities were mapped using GDSC2/ElasticNet modeling with cross-cohort replication.

**Results:** Our results define a reproducible ROS gradient, driven by effectors like *NQO1* and *SOD1,* characterizing ROS-high tumors as proliferative, epithelial and “immune-cold”. High residual ROS score was associated with an improved prognosis, regardless of TNM stage and age. Pharmacogenomic mapping revealed an overlapping sensitivity to mTOR inhibitors in ROS-high gastric cancer tumors which persisted after TME confounder adjustment.

**Conclusion:** The continuous ROS axis provides a functional readout of metabolic dependency that refines traditional anatomical staging. By identifying mTOR dependent cold tumors, our framework offers a precision strategy for immunotherapy-resistant patients like those affected by microsatellite-stable gastric cancer.

## BACKGROUND

Stomach adenocarcinoma (STAD) remains one of the leading causes of cancer-related deaths worldwide, ranking fifth in incidence and mortality on a global scale [1,2]. Poor prognosis in STAD is mainly due to its profound molecular and metabolic heterogeneity [3]. While the major classifications by The Cancer Genome Atlas (TCGA) and the Asian Cancer Research Group (ACRG) have provided an overview of the genomic landscape, translating these insights into precise therapeutic interventions has proven difficult. A crucial factor driving tumor heterogeneity is its intrinsic redox state[4,5]. While initially viewed merely as deleterious metabolic byproducts, reactive oxygen species (ROS) are now recognized as pivotal rheostats that regulate critical signaling pathways and dictate therapeutic response [6,7]. Existing paradigms often fail to capture the fluid, continuous nature of metabolic reprogramming during tumor evolution. Furthermore, the “intrinsic” ROS signature of a neoplastic cell is frequently masked by extrinsic tumor microenvironment (TME) factors, like immune infiltration and stromal remodeling [8,9]. Consequently, there is an urgent need for an analytical framework that can capture the continuous biomarkers and accurately reflect a tumor’s true oxidative stress-mediated vulnerability while itself remain independent of traditional clinical variables like TNM Stage.

In this study, we have addressed this gap by defining and validating a continuous ROS transcriptional axis using clinical data from independent gastric cancer cohorts. By utilizing the TCGA-STAD cohort [10], we have identified a coordinated gene program, driven by key redox regulators like *NQO1, SOD1* and *GPX4,* that defines a reproducible biological gradient. We have further demonstrated that this ROS Axis is not a cohort-specific artifact, but a conserved functional feature that can be replicated with high fidelity in the independent ACRG cohort [11]. We have also shown that this ROS Axis is a potent and independent prognostic factor for overall patient survival which remains significant even after adjusting for age and tumor stage – fundamental factors for clinical decision making [12]. Our finding shows that although higher intrinsic ROS scores provide a prognostic advantage, it uncovers a distinct therapeutic vulnerability. By applying a residual-based adjustment to separate the ROS signal from TME confounders, we have used an *in-silico* drug sensitivity screen via the Genomics of Drug Sensitivity in Cancer (GDSC) portal [13,14] to identify potential compounds that can target the high-ROS state. Our findings reveal a convergence of the therapeutic sensitivity on the mTOR signaling axis, as well as PI3K/AKT signaling, proteostasis via HSP90 and nucleotide metabolism. The convergence of the therapeutic sensitivity on both the PI3K/AKT/mTOR cascade and the HSP90 chaperone system in high-ROS gastric cancer patients suggests a unique metabolic vulnerability. Previous studies have highlighted the importance of inhibiting the HSP90 chaperone network [15] and have demonstrated how combinatorial mTOR inhibition can mitigate the compensatory effects of cellular stress response [16]. This presents an idea that ROS-high, immune-cold gastric tumors could be intrinsically dependent on the mTOR-HSP90 crosstalk and their therapeutic resistance could potentially be subdued by simultaneously disrupting both primary and adaptive resistance mechanisms. Based on our findings, we propose that the continuous ROS Axis identifies a unique biological gradient in stomach adenocarcinoma where high ROS has a more favorable baseline prognosis and is uniquely “primed” for targeted intervention. We also propose that a combinatorial treatment strategy utilizing Rapamycin (mTOR inhibitor) and Tanespimycin (HSP90 inhibitor) will improve clinical efficacy and patient prognosis in ROS-high stomach adenocarcinoma. We have provided a functional readout of metabolic dependency based on a ROS-driven framework that offers a strategy to revitalize potent agents – including clinically relevant AKT/mTOR and HSP90 inhibitors through patient stratification that complements and refines the traditional genomic and anatomical staging. Our results serve as a powerful predictor of ROS-driven drug sensitivities, offering a novel strategy for the clinical stratification of gastric cancer patients.

## METHODS

### Data Acquisition and Preprocessing

Gene expression profiles and corresponding clinical annotations for stomach adenocarcinoma (STAD) were obtained from The Cancer Genome Atlas (TCGA) through the ‘recount3’ resource. The TCGA-STAD dataset contained a total of 416 tumor samples. Raw RNA-seq count matrices were downloaded at the gene level and restricted to primary tumor samples based on curated TCGA sample annotations. Gene expression values were normalized to log2 counts per million (log2CPM) following library-size adjustment. To improve numerical stability and reduce noise in downstream matrix factorization analyses, genes exhibiting near-zero variance across samples were removed prior to model fitting. An independent gastric cancer validation cohort (n = 300) from the Asian Cancer Research Group (ACRG; GSE62254) was obtained from the Gene Expression Omnibus (GEO) [11]. This dataset served as an external cohort for validating transcriptional programs identified in the TCGA discovery cohort.

For pharmacological modeling, we utilized the Genomics of Drug Sensitivity in Cancer (GDSC2) resource. Pan-cancer cancer cell line gene expression profiles were integrated with their corresponding natural log-transformed half maximal inhibitory concentration values [ln(IC50)] for a large panel of anti-cancer compounds. These paired transcriptomic and drug-response datasets were used to train predictive models linking ROS-associated transcriptional programs to therapeutic sensitivity.

### Intrinsic ROS Gene Set Definition and Program Discovery

The Reactive Oxygen Species (ROS) pathway gene set was obtained from the Molecular Signatures Database (MSigDB v2025.1) [17,18]. This curated collection contained 48 ROS-associated genes (see Supplementary Fig. 1), all of which were present in the preprocessed TCGA-STAD expression dataset. Because simple unweighted average expression scores are often sensitive to biological noise and opposing pathway signals [19], Consensus Non-Negative Matrix Factorization (NMF) was applied to identify coordinated transcriptional programs within the ROS gene expression matrix. NMF decomposes the non-negative expression matrix X into two lower-rank matrices: where ‘W’ represents the sample-by-factor matrix describing the activity of each latent transcriptional program across samples, and ‘H’ represents the factor-by-gene matrix containing the gene-level loadings defining each program. Thus, the values in W quantify the activation strength of each transcriptional program in individual tumor samples. The decomposition follows: *X ≈ W × H*, where W represents sample-level program activity and H encodes gene-level contributions.

**Figure 1.**
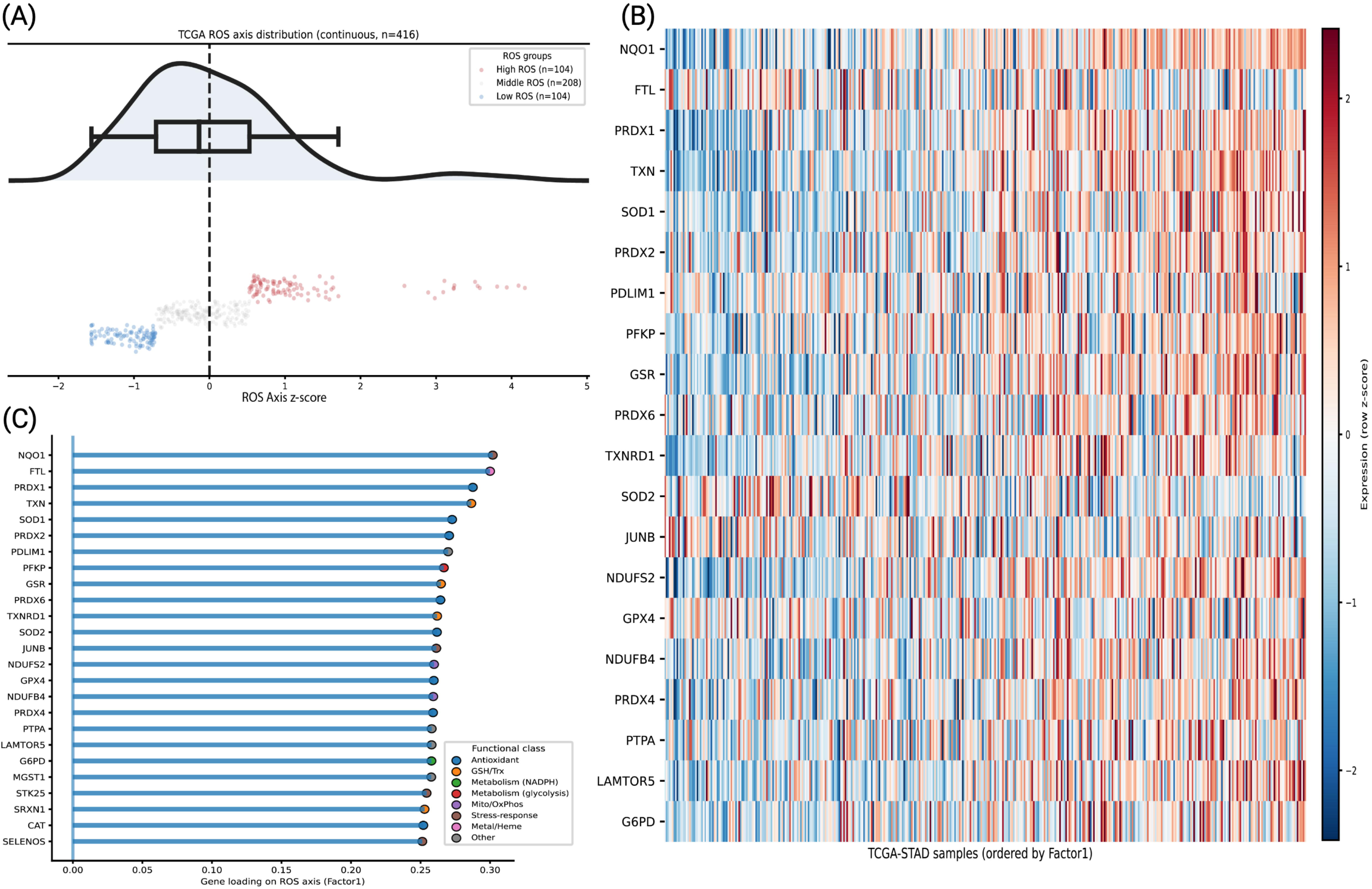
NMF deconvolution of bulk transcriptomes identifies a conserved ROS transcriptional axis in gastric adenocarcinoma. **(A)** Distribution of ROS axis scores (NMF Factor 1, z-standardised) across 416 TCGA-STAD tumours, shown as a kernel density estimate with overlaid boxplot. Samples were stratified into High (n=104), Middle (n=208), and Low (n=104) ROS tertiles for downstream analyses. **(B)** Expression heatmap of the 30 highest-loading Factor 1 genes across all samples ordered by ascending ROS score. Row-wise z-scored expression confirms progressive enrichment of redox transcripts across the axis. **(C)** Ranked NMF gene loadings colored by functional class, spanning antioxidant enzymes (NQO1, PRDX1, SOD1), glutathione/thioredoxin system members (TXN, GSR, SELENOS), NADPH metabolism (G6PD), and mitochondrial OXPHOS subunits (NDUFS2, NDUFB4), collectively defining a coordinated redox rheostat.

As log2CPM values may contain negative entries, a constant shift equal to the absolute minimum expression values plus a small pseudo-count (10^-6^) was applied to ensure strict non-negativity. NMF models were evaluated across ranks using 80 independent iterations per rank (NNDSVDa initialization, coordinate descent solver, Frobenius norm loss). The optimal rank of k was selected based on the cophenetic correlation coefficient of the consensus matrix and the silhouette score (see Supplementary Table 1). Ultimately, a two-program solution (k=2) was selected. NMF factors are permutation-invariant. Factor 1 was mathematically oriented, such that its sample weights positively correlated with the unweighted mean expression of the 48 ROS genes. To capture the entire biological spectrum of ROS activation, the sample-level activity score for Factor 1 was treated as a continuous variable in all downstream multivariable analyses.

### Independent Validation of the ROS Transcriptional Program

To verify the biological generalizability of the ROS-associated transcriptional program, the learned NMF model was projected onto an independent gastric cancer validation cohort (ACRG; GSE62254). A new dataset has its own cohort-specific technical biases or distinct population genetics. Hence, if we apply a *de novo* clustering or matrix factorization, it can produce disparate latent spaces. To rigorously test our specific model, we preserved the exact gene-weight matrix (H) derived from the final TCGA NMF model. Later, we computationally projected this model on the new ACRG cohort using Non-Negative Least Squares (NNLS) optimization. This algorithm calculates a ROS activity score for each ACRG patient (Sample-level), based on how well their tumor’s gene expression matches the TCGA-defined ROS profile. Prior to NNLS projection, the ACRG expression matrix was filtered to retain only the genes overlapping with the TCGA model, and a global minimum shift was applied to satisfy the non-negativity requirements of the algorithm. This approach allowed us to score the independent ACRG patients using the exact same transcriptional parameters established in the primary cohort.

### Tumor Microenvironment Deconvolution and Confounder Adjustment

To rigorously evaluate the influence of the tumor microenvironment (TME) on the discovered Reactive Oxygen Species (ROS) transcriptomic axis, canonical gene signatures were defined to represent distinct cellular compartments and states. Additionally, an NRF2 target signature of 20 canonical antioxidant response genes (NQO1, HMOX1, GCLC, etc.) was curated, and the mTOR signaling pathway was extracted from MSigDB Hallmark Collection (v2025.1). Gene expression was Z-score standardized row-wise prior to averaging. Bivariate correlations between the ROS axis and each TME score was scored by both Pearson’s r and Spearman’s ρ. Multivariable OLS regression with NRF2 transcriptional activity and mTOR pathway scores as dependent variables confirmed the independent contribution of the ROS axis after simultaneous covariate adjustment. To isolate the TME-independent component of NRF2 transcriptional activity, multivariable OLS regression was performed with the NRF2 module score as the dependent variable and the five TME scores as simultaneous covariates; the resulting residuals represent NRF2 activity orthogonal to TME composition and were subsequently correlated with the ROS axis. The association between the ROS axis and mTORC1 transcriptional activity was assessed by Pearson correlation in both the TCGA-STAD discovery cohort (n = 416) and the ACRG validation cohort (n = 300), with OLS regression lines and 95% confidence intervals displayed

For all downstream analyses, a confounder-residualised ROS score was derived by regressing the raw Factor 1 Z-score against all five TME scores in an OLS model and retaining the residuals. The pathway score Y was modeled as a function of the ROS axis and the suite of TME module scores:

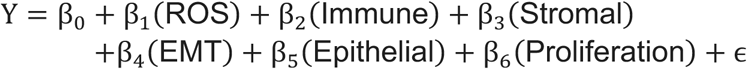

Models were fit utilizing the *statsmodels* framework in Python. For each parameter, the regression coefficient β, adjusted R^2^, and associated p-values were extracted to evaluate independent statistical significance and outcome variance explained by the model.

### Survival Analysis

Overall survival analyses were restricted to the TCGA-STAD cohort. Event status (1 = deceased) and survival time (days to death, or days to last follow-up for censored patients) were extracted from GDC clinical metadata. The TME-residualised ROS score was z-score standardized prior to all survival modelling.

Univariate survival analysis was performed by fitting a Cox Proportional Hazards model to the continuous residualised ROS score using the lifelines CoxPHFitter, with hazard ratios (HR), 95% confidence intervals, and Wald p-values reported. Kaplan–Meier curves were generated under two stratification schemes: median split and tertile split (top vs. bottom tertile), with between-group differences assessed by the log-rank test. A multivariate Cox model was subsequently fitted incorporating patient age at diagnosis, sex (male = 1), and pathological tumor stage (I–IV, ordinally encoded) to evaluate the independent prognostic contribution of the residualised ROS score after adjustment for established clinical confounders. Patients with missing values in any covariate were excluded from multivariate modelling.

### Pharmacogenomic Modelling of ROS-Associated Therapeutic Vulnerabilities

The 48 ROS pathway genes were used as the molecular feature space for drug sensitivity prediction. These were intersected with GDSC2 pan-cancer expression data, and the resulting feature matrix was merged with ln (IC₅₀) values via an inner join on cell line identifiers. A separate *ElasticNet* regression model was trained for each drug with ≥200 assayed cell lines (286 compounds total). Each pipeline comprised *StandardScaler* followed by *ElasticNetCV*, with hyperparameter optimization performed by 5-fold cross-validation over a grid of L1 ratios (0.1, 0.3, 0.5, 0.7, 0.9) and 30 logarithmically spaced α values (10^-3^ to 10^1^). Unbiased performance was assessed by out-of-fold predictions using *cross_val_predict,* with R^2^ and Spearman’s ρ computed against observed ln(IC_50_). Models with out-of-fold Spearman ρ ≥ 0.25 (n = 276 drugs) were deployed to impute drug sensitivity scores across TCGA-STAD and ACRG tumor samples, with missing ROS genes imputed as zero. Feature scaling was performed within cross-validation folds to prevent data leakage.

Drug-ROS associations were quantified by OLS regression of predicted ln(IC_50_) on the ROS axis in two stages: before and after TME residualisation. All p-values were corrected using the Benjamini–Hochberg procedure (FDR < 0.05). Association direction was classified as sensitivity (β < 0) or resistance (β > 0), and cross-cohort replication was defined as FDR-significant associations with concordant effect direction in both TCGA and ACRG after adjustment.

### Statistical Analysis and Software

All analyses were performed in Python 3.11 (*pandas, NumPy, SciPy, statsmodels, scikit-learn, lifelines, joblib*). Figures were generated with *Plotly* and exported via *Kaleido*.

## RESULTS

### Unsupervised decomposition defines a continuous and multi-functional transcriptomic gradient of oxidative stress in gastric cancer

Given the pivotal influence of reactive oxygen species (ROS) on the progression of tumors and their susceptibility to treatment, our initial investigation focused on whether oxidative stress in gastric cancer manifests as a simple binary condition or exists as a continuous biological gradient.

To define the transcriptional architecture, we used non-negative matrix factorization (NMF), an unsupervised, pathway-restricted decomposition of ROS-associated genes to the TCGA-STAD cohort. Rather than discretizing tumors into rigid and artificial subtypes, this approach helps us to model ROS activity as a fluid and continuous biological state. Consensus clustering across multiple ranks (k=2 to 6) identified k=2 as the optimal factorization rank based on peak stability metrics (cophenetic correlation = 1.0; silhouette score = 1.0) [see Supplementary Table 1]. To biologically orient the model, we correlated the derived latent components against a baseline ROS anchor signature. Factor 1 exhibited a robust and highly significant positive concordance(0.30; p< 0.0001, see Supplementary Table 2) and was consequently selected to serve as our continuous transcriptomic score for the ROS program. The distribution of this ROS axis (or factor 1) is represented in the form of low (bottom quartile), middle (interquartile range), and high (top quartile) ROS categories (dot plot) (Fig.1A) for visualization; the underlying signal varies continuously across the cohort (distribution plot). When TCGA-STAD samples were rank-ordered by their ROS axis score, we observed a distinct, continuous expression gradient of ROS-associated transcripts, further supporting the continuous nature of this biological axis (Fig. 1B).

Examination of gene loading (the H matrix) determined the biological drivers of this program. The highest-weighted genes driving the ROS program included major cytoprotective and detoxification effectors such as *NQO1, FTL, PRDX1* and *TXN*. Notably, the loading structure spans multiple functional classes like antioxidant systems, metabolic pathways and mitochondrial processes. Hence, ROS axis represents an integrated oxidative stress program rather than just a single pathway.

Together, these findings establish this ROS axis as a biologically coherent, continuous transcriptional axis of oxidative stress in gastric cancer. **A critical question, however, is whether this program is specific to the discovery cohort or reflects a conserved feature of stomach adenocarcinoma.**

### The ROS axis represents a conserved biological dimension reproducible across an independent gastric cancer cohort

To test the generalizability of the ROS axis, we projected the TCGA-derived ROS basis into an independent gastric cancer cohort (ACRG) of 300 patients, using a fixed gene loading matrix and Non-Negative Least Squares (NNLS). Projected ROS scores again followed a continuous distribution, mirroring a similar coordinated pattern like the discovery set (Fig. 2A). ACRG and the H matrix have 46 genes (out of 48) in common (see Supplementary Fig. 1), and a correlation value between the ROS axis and the ACRG corescore was found to be ρ= 0.63351 and p=4.572e^-35^. Rank ordering the ACRG samples by the projected score recapitulated the continuous expression gradient of the core ROS effectors (Fig.2B), showing cross-cohort stability of the signature.

**Figure 2.**
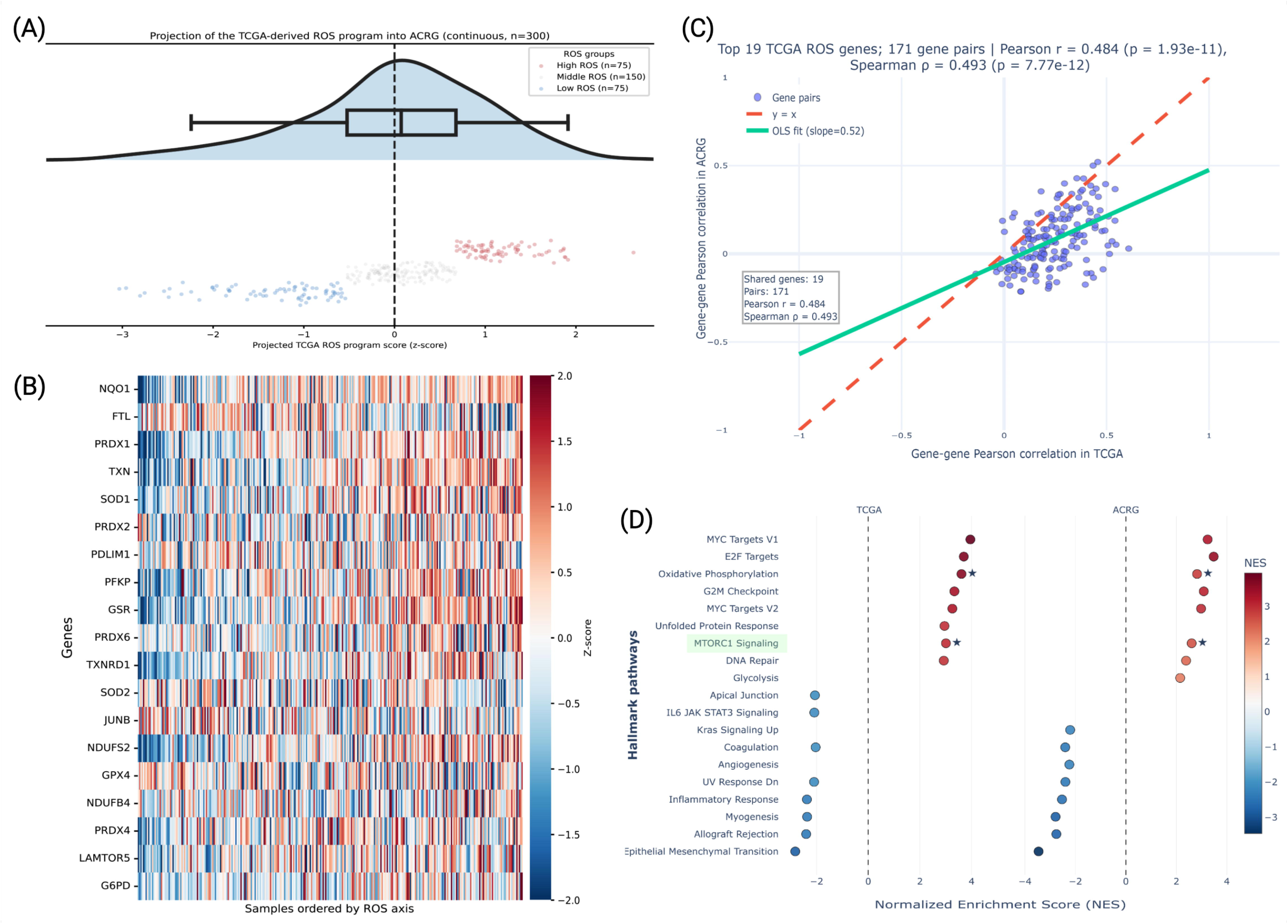
Cross-cohort projection validates the ROS transcriptional program in the independent ACRG cohort. **(A)** Distribution of TCGA-derived ROS scores projected onto 300 ACRG tumors (z-standardised), stratified into High (n=75), Middle (n=150), and Low (n=75) ROS tertiles. The unimodal distribution confirms stable program projection across independent platforms. **(B)** Expression heatmap of the 19 shared ROS signature genes across ACRG samples ordered by ascending projected score, recapitulating the coherent redox expression gradient observed in the discovery cohort. **(C)** Gene-pair correlation structure of the ROS program is preserved across cohorts. Pairwise Pearson correlations among the 19 shared genes (171 pairs) in TCGA and ACRG show significant concordance (Pearson r = 0.484, p = 1.93×10^-11^; Spearman ρ = 0.493, p = 7.77×10^-12^), with an OLS slope of 0.52 indicating moderate attenuation consistent with platform differences. **(D)** GSEA Hallmark pathway enrichment of the ROS axis in both cohorts. mTORC1 Signaling (highlighted) and Oxidative Phosphorylation are consistently and significantly enriched (★) across TCGA and ACRG, while immune and stromal program (IL6-JAK-STAT3, EMT, Inflammatory Response) are negatively associated, motivating downstream TME confounder adjustment.

To further estimate whether the internal structure of the ROS program is conserved or not, we examined the gene-gene correlation pattern among the top 19 genes between TCGA and ACRG (Pearson r = 0.484; Spearman ρ = 0.493; Fig. 2C). This also concludes that the transcriptional relation defining the ROS program is stable across datasets.

At the pathway level, the ROS axis showed consistent association with key oncogenic and metabolic processes in both the cohorts. Oxidative phosphorylation, MYC targets and mTORC1 signaling were positively enriched along the ROS axis. Conversely, low ROS scores were associated with epithelial-mesenchymal transition, inflammatory response, and angiogenesis. The concordance of these enrichments across TCGA and ACRG reinforces that the ROS axis captures a reproducible biological state with shared downstream consequences.

Ultimately, a successful projection and network-level replication in the ACRG cohort confirms that the ROS axis represents a fundamental dimension of gastric cancer biology. Having established the ROS axis as a conserved transcriptional program, we next asked **how it relates to the cellular phenotype and the tumor microenvironment.**

### The ROS axis identifies an immune-cold, proliferative phenotype driven by intrinsic NRF2 and mTOR signaling

If the ROS program reflects a fundamental axis between two gastric cancer cohorts, does it reflect intrinsic tumor biology, or is it driven by extrinsic factors like immune infiltration, stromal composition, or epithelial state?

Across the TCGA cohort, the ROS program showed a significant association with multiple tumor composition and state-related features (Fig.3A). Specifically, high ROS tumors exhibited elevated epithelial identity (Pearson *r* = 0.46, *P* = 1.1 × 10^-22^) and proliferative capacity (Pearson *r* = 0.43, *P* = 6.0 × 10^-20^), recapitulating the coordinated enrichment of MYC targets and cell cycle signaling programs observed in the preceding analysis. Conversely, ROS-high tumors showed lower immune infiltration (*r* = −0.51, *P* = 5.9 × 10^-29^), stromal/ECM content (*r* = −0.44, *P* = 6.0 × 10^-21^), and EMT features (*r* = −0.54, *P* = 4.8 × 10^-33^). These associations replicated in ACRG (n = 300; see Supplementary Fig. 2), with comparable or stronger effect sizes for epithelial identity (*r* = 0.61), proliferation (*r* = 0.56), EMT (*r* = −0.61), stromal/ECM (*r* = −0.59), and immune infiltration (*r* = −0.49). It directs towards the embedded nature of the ROS axis within broader tumor architectural and phenotypic variation. These strong bivariate associations confirmed that bulk-tissue composition is a strong confounder, where ROS-high tumors are highly proliferative, “immune-cold” and epithelial, whereas ROS-low tumors are stroma-rich.

**Figure 3.**
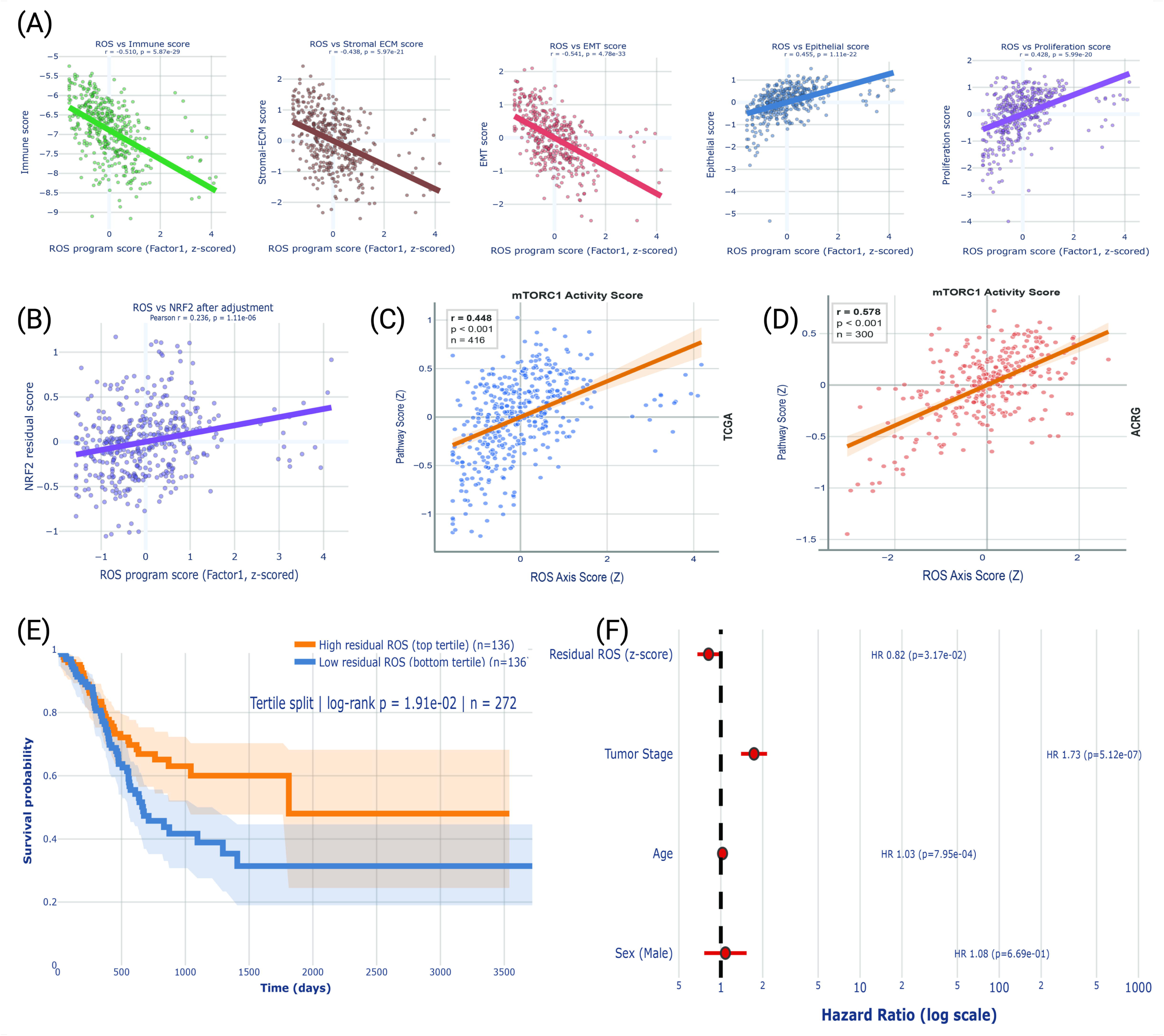
TME confounder adjustment unmasks an independent ROS signal with prognostic significance. **(A)** Pearson correlations between the raw ROS axis score and five TME program scores – Immune (r = −0.510), Stromal-ECM (r = −0.438), EMT (r = −0.541), Epithelial (r = 0.455), and Proliferation (r = 0.428) – confirming substantial confounding of the bulk ROS signal by microenvironmental composition. **(B)** Following multivariable OLS residualisation against all five TME covariates, the adjusted ROS score retains a significant positive association with NRF2 transcriptional activity (r = 0.236, p = 1.11×10^-6^), confirming that NRF2 pathway engagement is intrinsic to the tumour-cell ROS program. **(C-D)** The ROS axis score is positively correlated with mTORC1 activity score in both TCGA (r = 0.448, p < 0.001, n = 416) and ACRG (r = 0.578, p < 0.001, n = 300), demonstrating that the ROS-mTORC1 co-activation relationship is robust and cohort-independent. **(E)** Kaplan–Meier analysis of overall survival stratified by residual ROS tertile (n = 272; top vs. bottom, n = 136 each). High residual ROS is associated with significantly improved survival (log-rank p = 1.91×10^-2^), consistent with an adaptive NRF2-driven antioxidant phenotype. **(F)** Multivariable Cox regression confirms residual ROS score as an independent prognostic factor (HR = 0.82, p = 3.17×10^-2^), with tumour stage (HR = 1.73, p = 5.12×10^-7^) and age (HR = 1.03, p = 7.95×10^-4^) behaving as expected covariates. Sex was not independently prognostic (HR = 1.08, p = 0.669).

The pathway analysis in Figure 2D implicated mTORC1 signaling as enriched in ROS-high tumors. Because mTORC1 lies downstream of PI3K/AKT and is regulated in part by NRF2-mediated antioxidant signaling [20,21], we asked whether these pathway associations persist after accounting for shared variance with microenvironment features. Hence, we performed a multivariable regression analysis. We adjusted the ROS-program scores for EMT, Epithelial, and Immune infiltration metrics, thereby creating the residual transcriptomic variance attributable solely to the ROS axis. Even after this stringent adjustment, residual NRF2 and mTORC1 activity scores remained positively correlated with the ROS axis in TCGA (NRF2: r = 0.24, P = 1.1 × 10^-6^; mTORC1: r = 0.45, p < 0.001; Fig. 3B-C). This association replicated in the independent ACRG cohort, where the ROS axis score remained positively correlated with mTORC1 activity with a stronger effect size (r = 0.58, p < 0.001, n = 300; Fig. 3D). These findings indicate that the coupling between the ROS axis and NRF2/mTOR signaling is not a by-product of tumor composition but reflects intrinsic pathway co-regulation.

As the residual ROS signal captures intrinsic tumor biology, which is independent of microenvironmental confounders, we next evaluated the clinical relevance of this signal. Here, we performed survival analysis using confounder-adjusted ROS scores to ensure that any observed association reflects only the ROS axis itself rather than underlying differences in immune infiltration, stromal content, or epithelial state (log-rank *P* = 0.019; n = 136 per group; Fig. 3E). Patients in the top tertile of the residual ROS scores showed significantly improved overall survival compared to those in the bottom tertile. This finding may appear to be counterintuitive given the proliferative and immune-cold nature of ROS-high tumors. But if we consider the obtained features of ROS-low tumors, i.e. rich stroma, high EMT and inflammation, these are often consistent with poorer outcomes in gastrointestinal malignancies. Multivariable Cox regression confirmed that residual ROS score remained an independent predictor of survival after adjusting for tumor stage, age, and sex (HR = 0.82 per standard deviation increase, 95% CI: 0.69-0.97, *P* = 0.017; Fig. 3F). Tumor stage (HR = 1.71, *p* = 5.1 × 10⁻⁷) and age (HR = 1.03 per year, *p* = 7.95 × 10^-4^) were also significant. This is a result independent of sex (*p* = 0.67). (see Supplementary Table 4)

In summary, while the bulk ROS signature is heavily influenced by proliferative epithelial cells, multivariable adjustment confirms that it reflects intrinsic oxidative stress biology (*NRF2*) coupled with distinct oncogenic signaling through the mTOR pathway. Importantly, this confounder-adjusted ROS axis independently predicts patient survival, reinforcing its clinical relevance. These microenvironment and pathway associations raise a natural question: **can the ROS axis identify therapeutic vulnerabilities?**

### Cross-cohort mapping of the ROS axis reveals a conserved therapeutic sensitivity to mTOR inhibition in gastric cancer

Having established the ROS transcriptional program as a reproducible and biologically grounded axis, we next asked our final translational question of this study: Does this microenvironment-independent transcriptomic state predict actionable pharmacological vulnerabilities? To address this, we mapped the therapeutic landscape of the ROS axis using an ElasticNet model trained on the GDSC2 database to predict in silico drug sensitivities (LN IC50) for both TCGA and ACRG cohorts. We correlated ROS scores with imputed drug sensitivity (AUC) for hundreds of compounds, both before and after regressing out immune, stromal, EMT, epithelial, and proliferation signatures.

In both TCGA and ACRG, most drug-ROS associations were attenuated after confounder adjustment, indicating that many apparent sensitivities reflect tumor composition rather than intrinsic ROS biology (Fig. 4A). In ACRG, the number of significant sensitivity associations dropped from 73 to 46 after adjustment, and in TCGA, from 231 to 12 (Fig.4B). Resistance associations showed a similar pattern. This filtering step is crucial for this study, as it helps in recognizing drugs whose efficacy tracks with the ROS program itself from those confounded by stromal or immune content.

**Figure 4.**
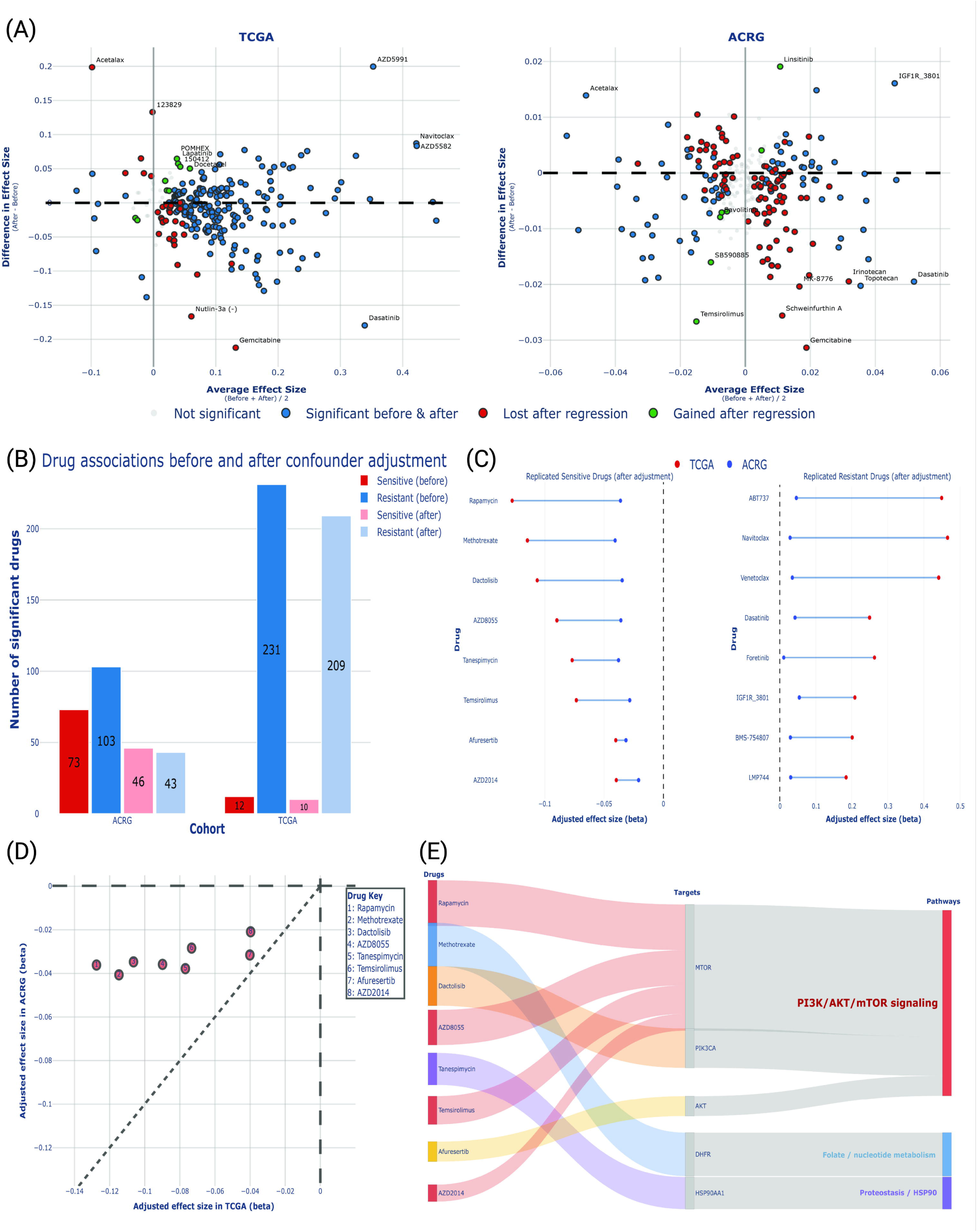
TME-adjusted ROS score identifies reproducible drug sensitivity associations across independent cohorts. **(A)** MA plots depicting drug–ROS association effect sizes before and after TME confounder adjustment in TCGA (left) and ACRG (right). Points are colored by association status: significant in both unadjusted and adjusted models (blue), lost after adjustment (red), or newly gained after adjustment (green), illustrating that residualisation both removes spurious TME-driven associations and unmasks tumor-intrinsic drug sensitivities. **(B)** Bar chart quantifying the number of significant drug associations (FDR < 0.05) before and after confounder adjustment, stratified by direction. Adjustment substantially reduces the number of apparent sensitivities, retaining only the most robust signals and enriching for resistance-associated compounds, particularly in TCGA. **(C)** Cross-cohort replicated drug associations after adjustment. Eight drugs are consistently associated with sensitivity in High-ROS tumors across both TCGA and ACRG (left), led by PI3K/AKT/mTOR inhibitors (Rapamycin, Dactolisib, AZD8055, Temsirolimus, Afuresertib, AZD2014) alongside Methotrexate and Tanespimycin. Nine drugs show concordant resistance associations (right), including ABT737, Navitoclax, Venetoclax, and Dasatinib. **(D)** Scatter plot of adjusted effect sizes (beta coefficients) for the eight replicated sensitivity-associated drugs in TCGA vs. ACRG. All points fall in the negative-negative quadrant with near-diagonal positioning, confirming directional and quantitative concordance of the ROS-drug relationship across platforms. **(E)** Sankey diagram mapping replicated sensitive drugs to their molecular targets and downstream pathways. The dominant convergence onto PI3K/AKT/mTOR signaling – through MTOR, PIK3CA, and AKT – across structurally distinct compounds establishes pathway-level rather than target-specific vulnerability in High-ROS gastric tumors.

To ensure maximal translational relevance, we strictly filtered for cross-cohort replication, selecting only those compounds that maintained significant association with the residual ROS axis in both TCGA and ACRG after the confounder adjustment. The stringent approach yielded a highly robust set of replicated sensitive (negative Beta values) and resistant (positive beta values) drugs (Fig. 4C). Interestingly, tumors with high intrinsic ROS activity showed striking resistance to BCL-2/BCL-XL family inhibitors (Venetoclax, Navitoclax, ABT737) and selective kinase inhibitors (Dasatinib, Foretinib, BMS-754807, LMP744). Ultimately, a refined subset of eight pharmacological compounds demonstrated robust and replicated sensitivity in tumors characterized by high intrinsic ROS activity: Rapamycin, Methotrexate, Dactolisib, AZD8055, Tanespimycin, Temsirolimus, Afuresertib, and AZD2014 or Vistusertib (Fig. 4C). Adjusted effect sizes for these drugs were directionally consistent across TCGA and ACRG, with all eight falling in the lower-left quadrant of a cross-cohort comparison plot (Fig.4D).

To explore the association of the sensitive drugs with specific pathways, we mapped the eight replicated sensitivity hits to their canonical targets and upstream pathways (Fig.5). Prominently, six out of those eight drugs directly target mTOR or its upstream PI3K/AKT signaling: Rapamycin, Temsirolimus, AZD8055 and AZD2014 or Vistusertib inhibit mTOR [22–25]; Dactolisib is a dual PI3K/mTOR inhibitor [26]; and Afuresertib targets AKT [27]. The remaining two drugs act on related vulnerabilities; Methotrexate inhibits DHFR [28] and Tanespimycin targets HSP90, a chaperone required for the stability of multiple oncogenic kinases [29]. This convergence reinforces the pathway-level findings from Figure 3.

The results collectively demonstrate that the ROS transcriptional program is not only biologically coherent but also functionally predictive of the therapeutic response. It works as an identifier of a reproducible and pathway-convergent vulnerability revolving around the mTOR signaling.

## DISCUSSION

Imbalanced redox state creates a high risk of generating higher level of reactive oxygen species in cancer cells that can modulate gene expression and modulate existing cellular programs [30]. Hence, it is quite necessary to characterize the oxidative stress in cancer biology. Discrete molecular subtypes or a binary stratification process imposes artificial boundaries by its biochemistry, which often is a graded cellular response [31,32]. Our study takes a different approach. We defined oxidative stress not as a categorial label but as a continuous transcriptomic gradient (Fig.1A) using NMF to ROS-associated transcripts. A single latent axis has finally been derived which captures the coordinated activity of cytoprotective effectors (*NQO1, PRDX1, TXN, FTL* [33–36]) alongside metabolic and mitochondrial programs (Fig.1B,C). This axis was not an artifact of the TCGA-STAD discovery cohort. Independent ACRG dataset-based projection via non-negative least squares reproduced both the continuous score distribution (Fig.2A) and the internal gene-gene correlation structure of the program (Pearson r = 0.484; Spearman ρ = 0.493 for top gene correlations across cohorts) (Fig.2B,C). We also investigated the pathway enrichment profile where oxidative phosphorylation, MYC targets, and mTORC1 signaling have been found at the high end. EMT, inflammatory response, and angiogenesis at the low end was likewise conserved (Fig.2D). These results establish the ROS axis as a stable, reproducible dimension of gastric cancer biology rather than a cohort-specific signature.

We recognized early that bulk-tissue ROS scores are heavily confounded by intra-tumor tissue heterogeneity. ROS-high tumors are predominantly epithelial, while ROS-low tumors are more stromal. We specifically addressed this by regressing out EMT, epithelial identity, and immune infiltration scores, isolating the residual transcriptomic variance attributable to the ROS axis alone (Fig.3A). Even after this stringent adjustment, the ROS axis maintained its positive correlation with *NRF2* (Fig.3B) and the mTOR pathway (Fig.3C,D) signatures. While NRF2 is a key mediator of cellular redox homeostasis, the crosstalk between ROS and the mTOR pathway helps increase the antioxidant capabilities of a cancer cell, mainly driven by an increased SOD1 activity [37]. High intrinsic ROS state further promotes the pivotal cellular signaling of the mTOR axis [38] to stabilize NRF2 [39] and provides just enough antioxidant support to maintain high rates of malignant growth and proliferation [38,40]. The coupling described in this study between oxidative stress and mTOR signaling is not a compositional artifact but is a genuine intrinsic co-regulation. This finding is consistent with the known crosstalk between NRF2 driven antioxidant programs and mTOR dependent metabolic reprogramming [41–43].

A central finding of this study may come off as an apparent paradox in the survival. Patients with high intrinsic ROS activity (after adjustment), showed significantly better survival than those with low ROS scores (log-rank P = 0.019) (Fig.3E). At first glance, this seems inconsistent with the hyper-proliferative, *MYC*-driven phenotype of ROS high tumors. However, the comparison becomes coherent when we consider what defines the opposite end of the spectrum, where ROS-low gastric cancers in the analysis are stroma-rich, EMT-high and inflamed. Such a profile has been repeatedly associated with chemoresistance, peritoneal dissemination and dismal prognosis in gastrointestinal malignancies [44–46]. The mesenchymal phenotype confers treatment difficulties through multiple mechanisms, like reduced drug perfusion through dense ECM, activation of survival signaling via integrin-mediated adhesion and intrinsic resistance of apoptosis [47–49]. On the other hand, the epithelial-rich proliferative ROS-high tumors, although biologically aggressive at a cellular level, may have retained greater level of sensitivity towards standard cytotoxic regimes. This phenomenon mostly happens due to their high replicative rate and oncogene addiction-dependent apoptosis [50].

Mapping the pharmacological landscape of the ROS-axis via GDSC2 modeling revealed the absolute necessity of the microenvironmental deconvolution. After adjustment for confounders, the number of significant drugs (from 231 to 12, in TCGA) confirmed that the most bulk-tissue sensitivities merely reflect stromal or immune content. What survived this filtering and replicated across both the TCGA and ACRG cohorts, was a strikingly convergent set of vulnerabilities (Fig.4A,B). Six of the eight replicated sensitive hits – rapamycin, Temsirolimus, AZD2014 (Vistusertib), Dactolisib, AZD8055, and Afuresertib directly target the mTOR cascade either by directly inhibiting the mTORC1/2 complex or its upstream regulators [22–27]. The rest of the two sensitive compounds, Methotrexate and Tanespimycin, target adjacent nodes (Fig.4C-E). Methotrexate, a DHFR inhibitor, affects the carbon metabolism that feeds into mTOR-regulated biosynthetic demands [51,52]. Tanespimycin is a HSP90 inhibitor, and this chaperone is necessary for the folding and the stability of multiple oncogenic kinases, including AKT itself [53,54].

The ROS-high and potentially immune-cold population mentioned in our study is defined by a distinct metabolic dependency, specifically an addiction to the mTOR signaling pathway. These metabolic features often fall into the category of microsatellite stable (MSS) gastric tumors which are notoriously resistant to immune checkpoint inhibitors (ICI) [55–57]. Part of their resistance is driven by an enriched mTOR signaling which not only acts as a critical survival engine to buffer excessive oxidative stress but is also a known driver of the “cold” TME [58,59]. The sensitive drug hits identified in our analysis offer a potential precision therapeutic strategy for these ICI resistant patients. By disrupting the mTOR cascade, these agents may simultaneously collapse the tumor’s antioxidant defense and potentially “warm up” the TME by reversing pathway-mediated immune suppression. Moreover, the observed co-sensitivity to both mTOR and HSP90 inhibitors (Fig.4E) in the ROS-high population suggests a profound therapeutic vulnerability. Inhibition of HSP90 typically triggers compensatory cellular heat shock response mechanisms which attenuate the cytotoxic effects of the treatment. However, evidence suggests that this protective response is critically dependent on an mTOR mediated transcriptional activity [16]. By employing a combinatorial strategy to inhibit mTOR (via Rapamycin) and HSP90 (via Tanespimycin) it could be possible to suppress the nuclear translocation of HSF1, thereby preventing an adaptive stress response in ROS-high gastric cancer. This dual blockade will effectively collapse the tumor’s primary survival mechanism while simultaneously removing the compensatory heat shock response activation [15,16]. Such combinations have been previously tested in other cancers [60] but have not been reported in gastric cancer. Collectively, our findings suggest that for patients with a high ROS signature, the path to clinical efficacy lies in the targeted inhibition of metabolic axes like the mTOR signaling, providing a roadmap to categorize patients into trials where these specific inhibitors have the highest likelihood of success.

Certain limitations of the current investigation necessitate careful consideration. Primarily, our findings are predicated upon bulk transcriptomic datasets-specifically RNA-sequencing for the TCGA-STAD discovery cohort and microarray platforms for the ACRG validation cohort. While our rigorous multivariable adjustment for microenvironmental signatures mathematically mitigates compositional confounding, it lacks the resolution to fully delineate the cellular provenance of the ROS signal. Future integration of single-cell or spatial transcriptomic profiling remains essential to determine whether this ROS program is an intrinsic feature of neoplastic epithelial cells, a characteristic of specific stromal compartments, or an emergent property of their complex interaction. Furthermore, our pharmacogenomic predictions rely on *in-silico* imputation derived from cell-line-trained models (GDSC2/ElasticNet) rather than direct pharmacological measurements in patient-derived systems. Although cross-cohort replication across disparate expression profiling technologies bolsters confidence in these associations, prospective validation in patient-derived organoids or xenograft models is imperative before clinical implementation. Finally, while our survival analysis accounts for major clinical covariates and microenvironmental confounders, its retrospective nature necessitates confirmation within independent prospective cohorts utilizing standardized treatment protocols.

In conclusion, by treating oxidative stress as a continuous transcriptomic axis rather than a categorical label, and by rigorously removing microenvironment-driven variance, we have identified a biologically coherent and clinically actionable dimension of gastric cancer. This confounder-adjusted framework reveals that intrinsic ROS-high tumors are functionally dependent on mTOR signaling and provides a rational basis for patient stratification. Moreover, the obtained drug sensitivities offer a unique outlook on combinatorial therapies to improve the prognosis of ICI-resistant tumors. Implementing such a continuous transcriptomic strategy may offer a path to revive mTOR-targeted therapies in gastric oncology-not by deploying them broadly, but by directing them to the context of metabolic dependency in which they are most likely to succeed.

## Additional Information

## Acknowledgements

We are thankful to the KIIT School of Biotechnology, KIIT Deemed to be University and all the participants who made this study possible. Graphical figures representing workflow and graphical abstract were created using BioRender (https://BioRender.com).

## Authors’ contributions

**R.R.** and **J.P.** conceptualized the study, performed the analyses, and drafted the original manuscript. **R.R.** developed the computational pipeline, conducted the formal statistical and pharmacogenomic analyses, and generated the visualizations. **J.P.** contributed to data validation and biological interpretation. **A.C.** assisted with data validation and critical review of the manuscript. **S.P.** and **T.P.** supervised the project, provided overarching intellectual direction, and rigorously edited the final manuscript. All authors read and approved the final manuscript.

## Ethics approval and consent to participate

Human participants or human tissues were not used for this study. The *in-silico* analyses on patient data were done using publicly available datasets from TCGA and ACRG.

## Consent for publication

No individual person’s data liable for consent was used for this study.

## Data availability

**1**. Public datasets analyzed in this study include TCGA-STAD (via the recount3 package), the ACRG validation cohort (GEO: GSE62254), and GDSC2 pharmacogenomic profiles (www.cancerrxgene.org).
**2**. All processed data, trained models, and numerical outputs required to reproduce this study have been deposited on Zenodo (https://doi.org/10.5281/zenodo.19534167). The Zenodo repository is under restricted access during peer review and will become fully public upon publication. Intermediate access requests may be directed to the corresponding authors.

## Funding information

The authors received no financial support for the research, authorship, and/or publication of this article.

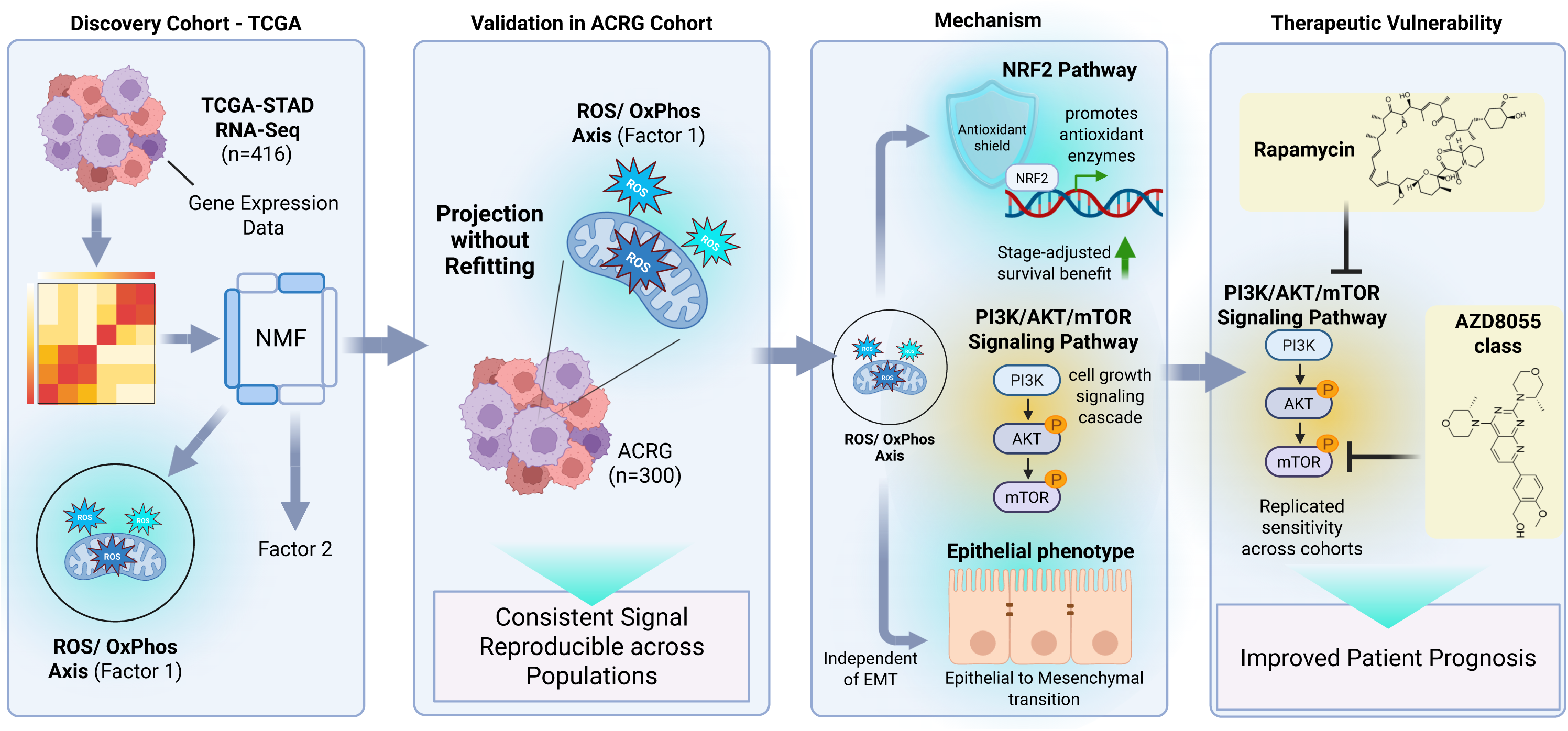

