## Supplementary Figures for "NMF Deconvolution of a High-ROS Transcriptional Program Uncovers mTOR-Dependent Therapeutic Sensitivity in Stomach Adenocarcinoma"

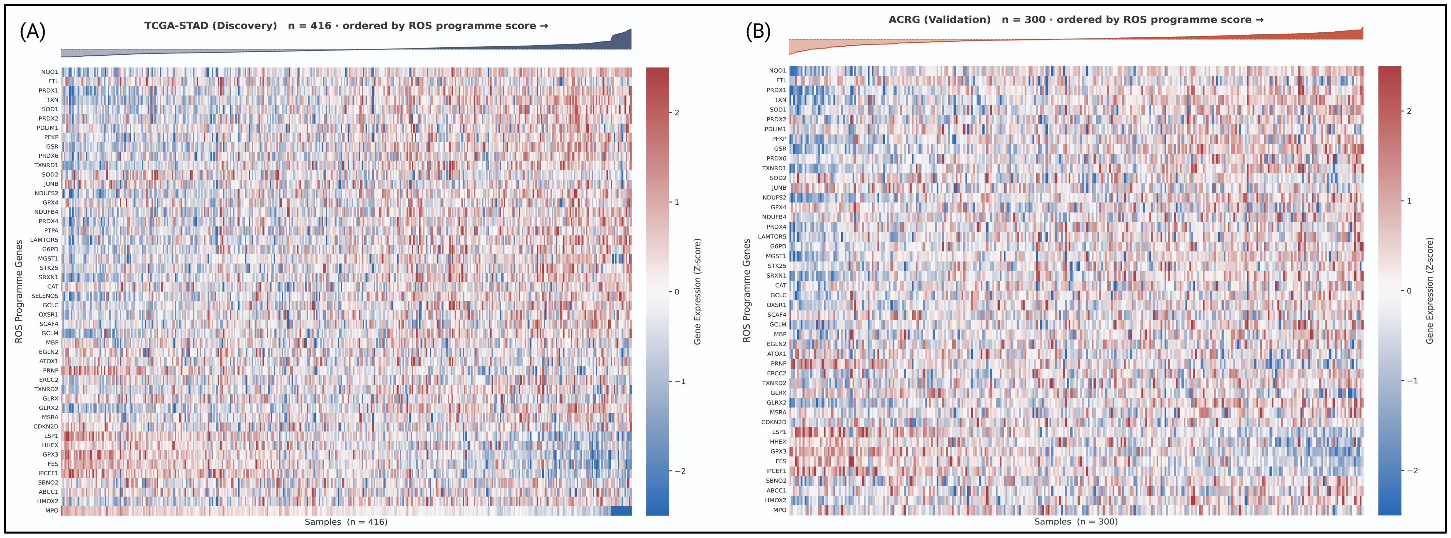


**Supplementary Figure 1. ROS gene expression gradient across cohorts.** Heatmaps of ROS pathway gene expression (z-score) in (A) TCGA-STAD (n = 416) and (B) ACRG (n = 300), with samples ordered by increasing ROS program score.

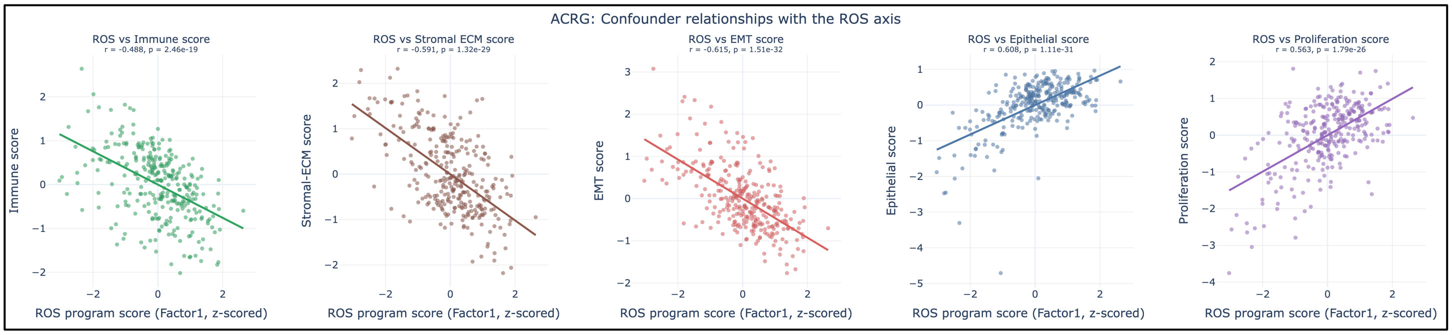


**Supplementary Figure 2. TME confounder relationships with the ROS transcriptional axis**. Pearson correlations between the projected ROS axis score (NMF Factor 1, z-standardised) and five TME programme scores in ACRG (n = 300). The ROS score has negative correlation with Immune ($r = -0.488$, $p = 2.46\times10^{-19}$), Stromal-ECM ($r = -0.591$, $p = 1.32\times10^{-29}$), and EMT ($r = -0.615$, $p = 1.51\times10^{-32}$) signatures, and positive correlation with Epithelial ($r = 0.608$, $p = 1.11\times10^{-31}$) and Proliferation ($r = 0.563$, $p = 1.79\times10^{-26}$) scores.

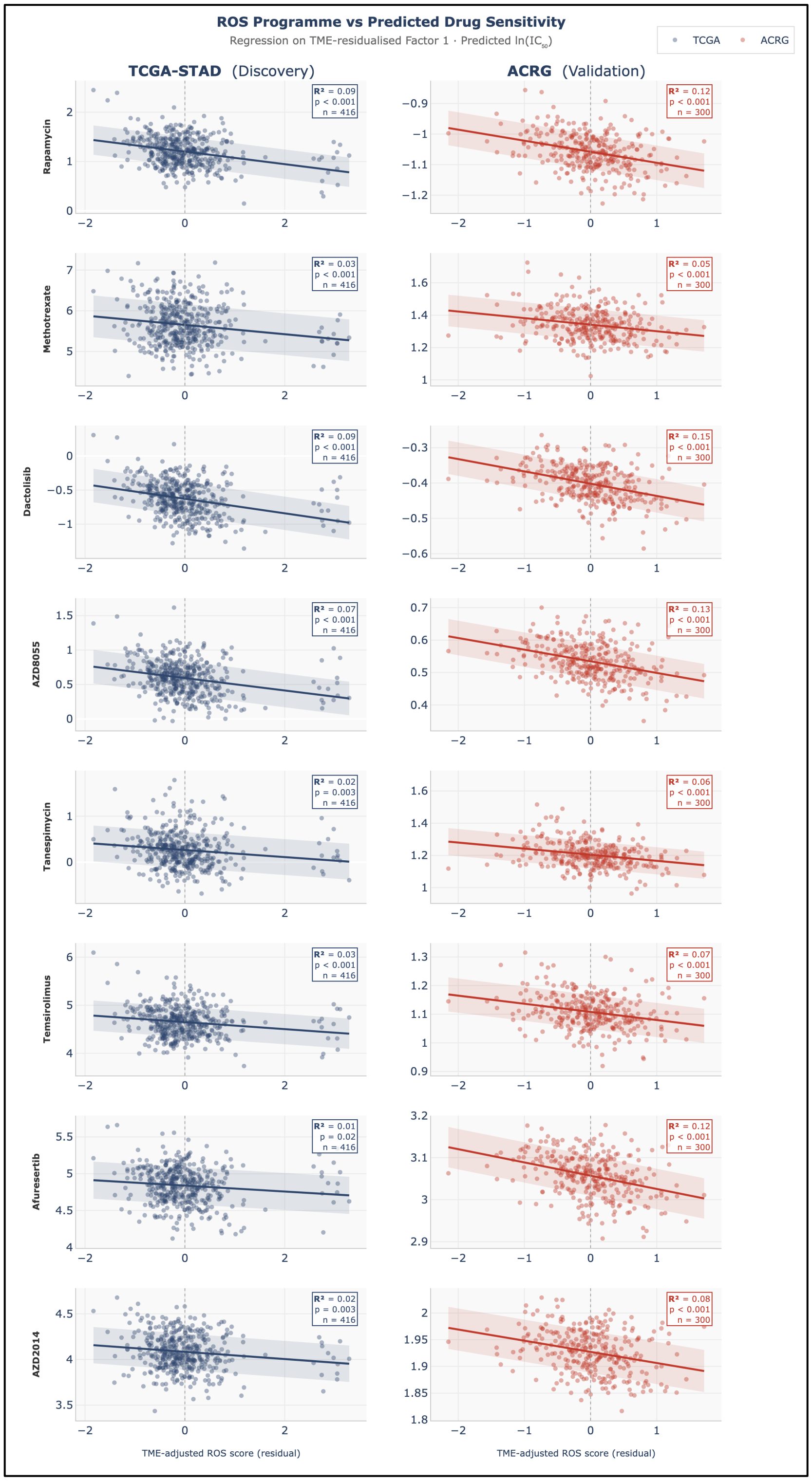


**Supplementary Figure 3. ROS program associates with predicted drug sensitivity across cohorts.** Scatter plots showing the relationship between TME-residualised ROS scores and predicted drug sensitivity ($\ln(IC_{50})$) for eight replicated drugs, both in TCGA (left, n = 416) and ACRG (right, n = 300). Each point represents a tumor sample; lines indicate linear regression with 95% confidence intervals. $R^2$ and p-values are shown in each panel. Higher ROS scores are consistently associated with lower predicted $\ln(IC_{50})$ (increased sensitivity) across both cohorts.
