## Supplementary Tables for "NMF Deconvolution of a High-ROS Transcriptional Program Uncovers mTOR-Dependent Therapeutic Sensitivity in Stomach Adenocarcinoma"

**Supplementary Table 1:** NMF rank selection metrics across candidate factorization ranks.

| k | cophenetic | silhouette | mean_ARI_vs_best | median_ARI_vs_best | best_seed | best_recon_error | n_runs |
| --- | --- | --- | --- | --- | --- | --- | --- |
| 2 | 1.0 | 1.0 | 1.0 | 1.0 | 28 | 0.7870993416764500 | 80 |
| 3 | 1.0 | 0.9951923076923080 | 1.0 | 1.0 | 75 | 0.7294072530422240 | 80 |
| 4 | 1.0 | 0.9975961538461540 | 1.0 | 1.0 | 20 | 0.6895405878984940 | 80 |
| 5 | 0.9985002807795260 | 0.9884944779015400 | 0.9693657990633260 | 0.9712795872309140 | 17 | 0.6527578116417890 | 80 |
| 6 | 0.999054875241513 | 0.9855518171442740 | 0.9599091044672520 | 0.9607660550471750 | 62 | 0.6152893445591860 | 80 |

**Supplementary Table 2:** Correlation of NMF factors with the ROS transcriptional anchor score.

| factor | spearman_rho_with_ROS_anchor | p_value |
| --- | --- | --- |
| Factor1 | 0.30022541267638900 | 4.11115512287476E-10 |
| Factor2 | -0.299902654336216 | 4.30075513823499E-10 |

**Supplementary Table 3:** Gene signatures used for tumor microenvironment and pathway scoring

| Signature | Genes |
| --- | --- |
| Immune | PTPRC, LST1, TYROBP, FCGR3A, CD3D, CD3E, TRAC, MS4A1, CD79A |
| Stromal/ECM | COL1A1, COL1A2, COL3A1, DCN, LUM, COL5A1, FN1, SPARC, ACTA2 |
| EMT | VIM, ZEB1, ZEB2, SNAI1, SNAI2, TWIST1, ITGA5, FN1 |
| Epithelial | EPCAM, CDH1, KRT8, KRT18, KRT19, MSLN, MUC1 |
| Proliferation | MKI67, TOP2A, HMGB2, UBE2C, CENPF, CCNB1, AURKB |
| NRF2 Targets | NQO1, HMOX1, GCLC, GCLM, SLC7A11, TXNRD1, SRXN1, GSR, PRDX1, PRDX6, FTL, FTH1, GPX2, GPX4, SOD1, ABCC1, ABCC2, UGT1A1, UGT1A6, ME1 |

**Supplementary Table 4:** Multivariable Cox proportional hazards model for overall survival in TCGA-STAD

| Covariate | Coeff. | Hazard_Ratio | se(coeff.) | P_Value | Coeff. lower 95% | Coeff. upper 95% |
| --- | --- | --- | --- | --- | --- | --- |
| ROS_resid_z | -0.199453047687379 | 0.8191786822435720 | 0.09378611790060670 | 0.03344669237481640 | -0.3832704610223960 | -0.015635634352362500 |
| Age | 0.028109676790381800 | 1.028508481744850 | 0.008564065899343160 | 0.001029706799639730 | 0.01132441606644160 | 0.04489493751432210 |
| Sex | 0.09973357644559840 | 1.1048765137313 | 0.1801773104514500 | 0.5799004306375540 | -0.2534074628705360 | 0.45287461576173200 |
| Stage | 0.5340339903863320 | 1.705799627254530 | 0.10948920031202300 | 1.07436780090596E-06 | 0.31943910107867500 | 0.7486288796939890 |
